# Association of Nirmatrelvir/Ritonavir Treatment with Long COVID Symptoms in an Online Cohort of Non-Hospitalized Individuals Experiencing Breakthrough SARS-CoV-2 Infection in the Omicron Era

**DOI:** 10.1101/2023.03.02.23286730

**Authors:** Matthew S. Durstenfeld, Michael J. Peluso, Feng Lin, Noah D. Peyser, Carmen Isasi, Thomas W. Carton, Timothy J. Henrich, Steven G. Deeks, Jeffrey E. Olgin, Mark J. Pletcher, Alexis L. Beatty, Gregory M. Marcus, Priscilla Y. Hsue

## Abstract

**Background:** Oral nirmatrelvir/ritonavir is a treatment for COVID-19, but whether treatment during the acute phase reduces the risk of developing Long COVID is unknown.

**Methods:** Using the Covid Citizen Science (CCS) online cohort, we surveyed individuals who reported their first SARS-CoV-2 positive test between March and August 2022 regarding Long COVID symptoms. We excluded those who were pregnant, unvaccinated, hospitalized for COVID-19, or received other antiviral therapy. The primary exposure was oral nirmatrelvir/ritonavir. The primary outcome was the presence of any Long COVID symptoms reported on cross-sectional surveys in November and December 2022. We used propensity-score models and inverse probability of treatment weighting to adjust for differences in treatment propensity. Our secondary question was whether symptom or test positivity rebound were associated with Long COVID.

**Results:** 4684 individuals met the eligibility criteria, of whom 988 (21.1%) were treated and 3696 (78.9%) were untreated; 353/988 (35.7%) treated and 1258/3696 (34.0%) untreated responded to the survey. Median age was 55 years and 66% were female. We did not identify an association between nirmatrelvir/ritonavir treatment and Long COVID symptoms (OR 1.15; 95%CI 0.80-1.64). Among n=666 treated with nirmatrelvir/ritonavir who responded who responded to questions about rebound, rebound symptoms or test positivity were not associated with Long COVID symptoms (OR 1.34; 95%CI 0.74-2.41; p=0.33).

**Conclusions:** Within this cohort, treatment with nirmatrelvir/ritonavir among vaccinated, non-hospitalized individuals was not associated with lower prevalence of Long COVID symptoms or severity of Long COVID. Experiencing rebound symptoms or test positivity is not strongly associated with developing Long COVID.

## Introduction

Symptoms after SARS-CoV-2 may persist as Long COVID, a type of post-acute sequalae of SARS-CoV-2 infection (PASC) defined as unexplained symptoms attributed to COVID-19 that are present at least 90 days following initial infection and persist for at least 2 months [1]. There are no treatments yet identified to reduce the risk of developing Long COVID. Vaccination reduces but does not eliminate the risk of Long COVID [2]. Higher acute viral load or prolonged viral shedding may be associated with increased risk of Long COVID [3, 4]. Recent studies suggest viral persistence in a subset of individuals with post-acute sequelae of COVID-19, including prolonged gastrointestinal shedding, ongoing Spike and Nucleocapsid antigen in neuronal and astrocytic exosomes, and evidence of persistent viral RNA or proteins in deep tissues [5-8]. Among high-risk non-hospitalized individuals with symptomatic COVID-19, nirmatrelvir, a novel oral orally administered severe acute respiratory syndrome coronavirus 2 main protease (M_pro_) inhibitor in combination with ritonavir reduces viral load and progression to severe disease [9]. Although anecdotal reports suggest that nirmatrelvir/ritonavir treatment during acute SARS-CoV-2 infection may improve Long COVID symptoms [10-12], this has not been replicated in an observational target-trial emulation which found no difference in the incidence of EHR-diagnosed post-COVID conditions after 30 days among those treated with nirmatrelvir/ritonavir [13]. However, use of EHR-based diagnosis of post-COVID conditions, which rely on patient reporting and clinician documentation of medical diagnoses, may not capture the possible treatment effect on Long COVID symptoms.

Therefore, the objectives of this study were to test the hypothesis that nirmatrelvir/ritonavir treatment during acute SARS-CoV-2 infection is associated with a reduction in patient-reported Long COVID symptoms at least 90 days after acute infection, and secondly whether patient-reported rebound of acute symptoms or rebound test-positivity after treatment with nirmatrelvir/ritonavir is associated with Long COVID symptoms. We hypothesized that treatment with nirmatrelvir/ritonavir would be associated with lower risk and rebound with higher risk of Long COVID symptoms.

## Methods

### Design, Setting, and Participants

The COVID-19 Citizen Science (CCS) Study is an online cohort study mostly of United States residents hosted on the Eureka Research Platform by the University of California, San Francisco [14]. Recruitment occurred through email invitations to existing participants in other Eureka studies, referrals from partner organizations, press releases, and word of mouth. To participate, individuals must be at least 18 years old, register for a Eureka Research Platform account with an Android, iPhone, or on the web-based study, agree to participate in English and provide electronic consent. After consenting, participants complete baseline surveys and are subsequently asked to complete regular surveys. This study included data collected from March 26, 2020 to December 22, 2022. For the primary analyses, we included individuals who reported their first positive SARS-CoV-2 test (antigen or PCR) in March 2022 through August 2022. Outcomes were ascertained only for those who responded to surveys about Long COVID symptoms sent out in November 2022 and December 2022. The primary outcome was patient-reported Long COVID symptoms at least 90 days after acute SARS-CoV-2 infection [1].

### Inclusion and Exclusion Criteria

We included CCS participants with first-reported positive SARS-CoV-2 antigen or PCR test starting in March 2022. Participants were excluded if they had ever reported a positive test for SARS-CoV-2 prior to March 2022. To ascertain the possible treatment effect of nirmatrelvir/ritonavir compared to no treatment, we excluded those who received other antiviral treatment (molnupiravir, remdesivir, and monoclonal antibodies). Due to very small numbers of individuals hospitalized for COVID-19 (n=1 in each group) and because >98% of individuals were vaccinated prior to SARS-CoV-2 infection we limited the primary analysis to non-hospitalized, vaccinated individuals. Finally, because no pregnant individuals were treated, we excluded individuals known to be pregnant from the control group.

### Exposures

The primary exposure was self-reported treatment with nirmatrelvir/ritonavir within 30 days of first reported positive SARS-CoV-2 antigen or PCR test on a weekly survey from March 2022-August 2022. Participants who self-reported taking nirmatrelvir/ritonavir were subsequently invited to respond to a survey in December 2022 to assess for rebound symptoms or test positivity. Rebound symptoms were defined as symptom worsening after initial improvement (“Did your COVID-related symptoms worsen after initially improving while taking or within a few days after completing Paxlovid?”). Rebound test positivity was defined as testing negative (“Did you ever test negative while taking Paxlovid or within 2 weeks after completing Paxlovid?”) and then positive on antigen test after completing treatment (“Did you subsequently test positive within two weeks of stopping Paxlovid?”).

### Outcomes

The primary outcome was ≥1 self-reported Long COVID symptom more than 90 days after first SARS-CoV-2 positive test on a cross-sectional survey distributed in November 2022 that asked about presence, duration, and severity of Long COVID symptoms using a non-validated instrument as we have previously published [15]. Symptoms queried included fatigue, shortness of breath, confusion, headache, altered taste and smell, joint pain, muscle aches, cough, chest pain, scratchy throat, nausea, vomiting, diarrhea, fever, chills, red or painful eyes, sore throat, and other.

### Other variables

Other variables were self-reported by participants including demographics, medical history, vaccine history, and lifestyle factors as we have previously reported [15]. Only body mass index had >1% of data missing (650 missing, 40%). For the propensity adjusted model, we imputed missing body mass index from age, sex, race/ethnicity, and past medical history.

### Statistical Analysis

The analysis plan was developed prior to data collection. First, we compared individuals who did and did not receive nirmatrelvir/ritonavir using Chi-squared tests for categorical variables and T-tests for continuous variables. We used logistic regression models adjusted first for age, sex, and time since COVID-19 diagnosis, and then with past medical history and substance use added. Next we modeled propensity to take nirmatrelvir/ritonavir using logistic regression models including all individuals who met the inclusion/exclusion criteria regardless of whether they responded to the cross-sectional Long COVID survey including age, sex, race, ethnicity, socioeconomic status, education, employment, past medical history, substance use, number of vaccines. After checking the Hosmer-Lemshow goodness of fit and balance of covariates by propensity score quintile, we used logistic regression models to model the association between use of nirmatrelvir/ritonavir and self-reported Long COVID among survey respondents adjusted for age, sex, time since SARS-CoV-2 test positivity, and the cubic spline of the propensity. As a sensitivity analysis, we repeated the analysis using inverse probability of treatment weighting, truncating extreme weights >95th percentile. For analyses of rebound, we used Fisher’s exact test for unadjusted analyses as we did not expect age or sex to be associated with rebound so we did not consider them to be likely to be confounders. Analyses were conducted in SAS version 9.4 and STATA 17.0. Sample size was not determined a priori.

### Results Reporting and Informed Consent

Results are reported in accordance with STROBE guidelines.[16] All participants provided digitally-signed informed consent. The study was reviewed and approved by the UCSF Institutional Review Board (#17-21879).

## Results

Within the CCS, 4684 individuals reported their first positive test for SARS-CoV-2 during the study period and met the eligibility criteria, of whom 988 (21.1%) were treated with nirmatrelvir/ritonavir and 3696 (78.9%) were not treated (Figure 1). As expected based on prescribing guidelines [17], individuals treated with nirmatrelvir/ritonavir were older, more likely to be male, and had more comorbidities including hypertension, diabetes, asthma, sleep apnea, and cardiovascular disease (Table 1). There were no differences in the number of vaccines received prior to infection or the mean number of days since infection. Among those eligible, 353/988 (35.7%) treated and 1258/3696 (34.0%) untreated individuals responded to the Long COVID cross-sectional survey. Demographics, comorbidities, and time since SARS-CoV-2 infection were similar among respondents to the Long COVID survey compared to the overall cohort (Appendix Table 1).

**Figure 1.**
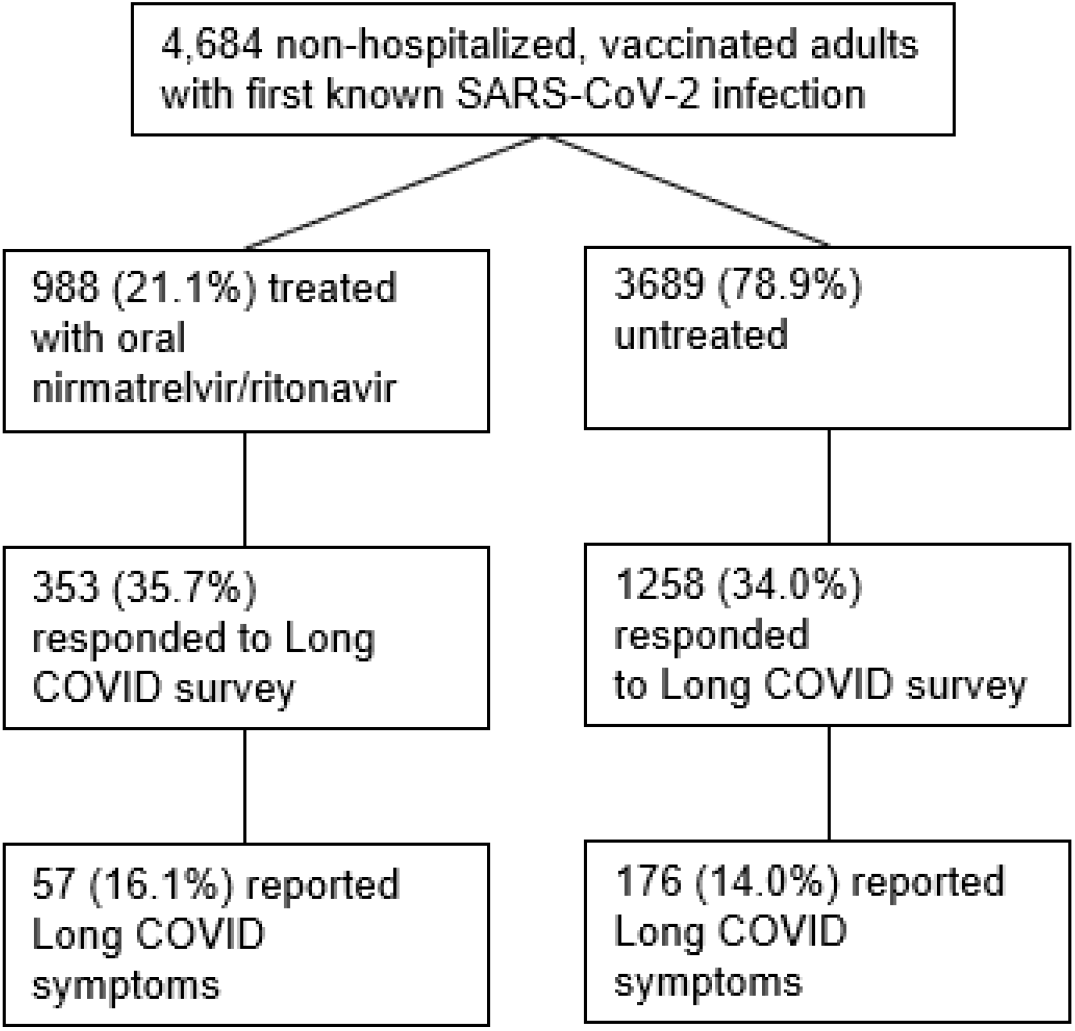
Participant Flow Diagram. Flow diagram for participants included in the primary analysis.

**Table 1.**
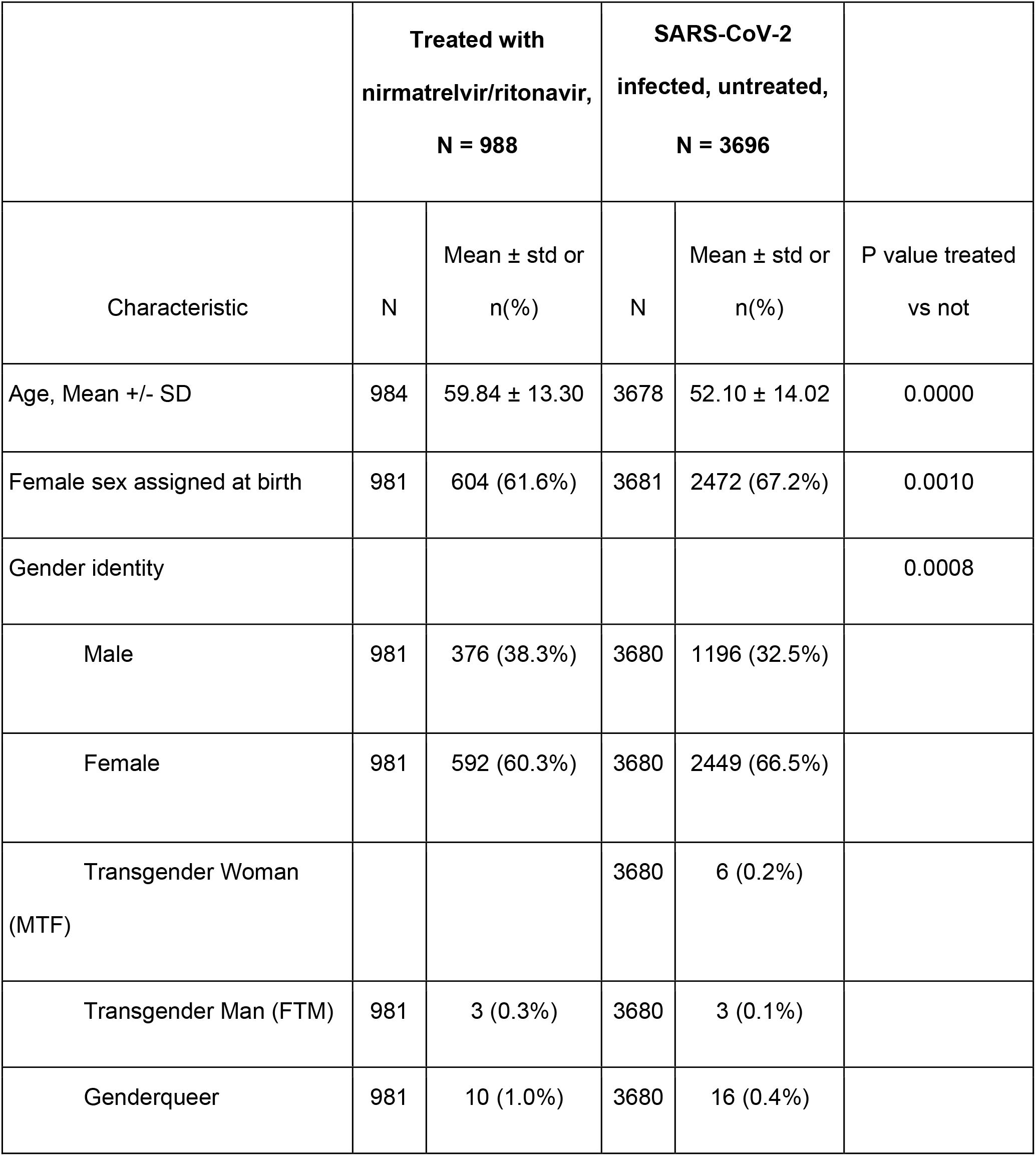
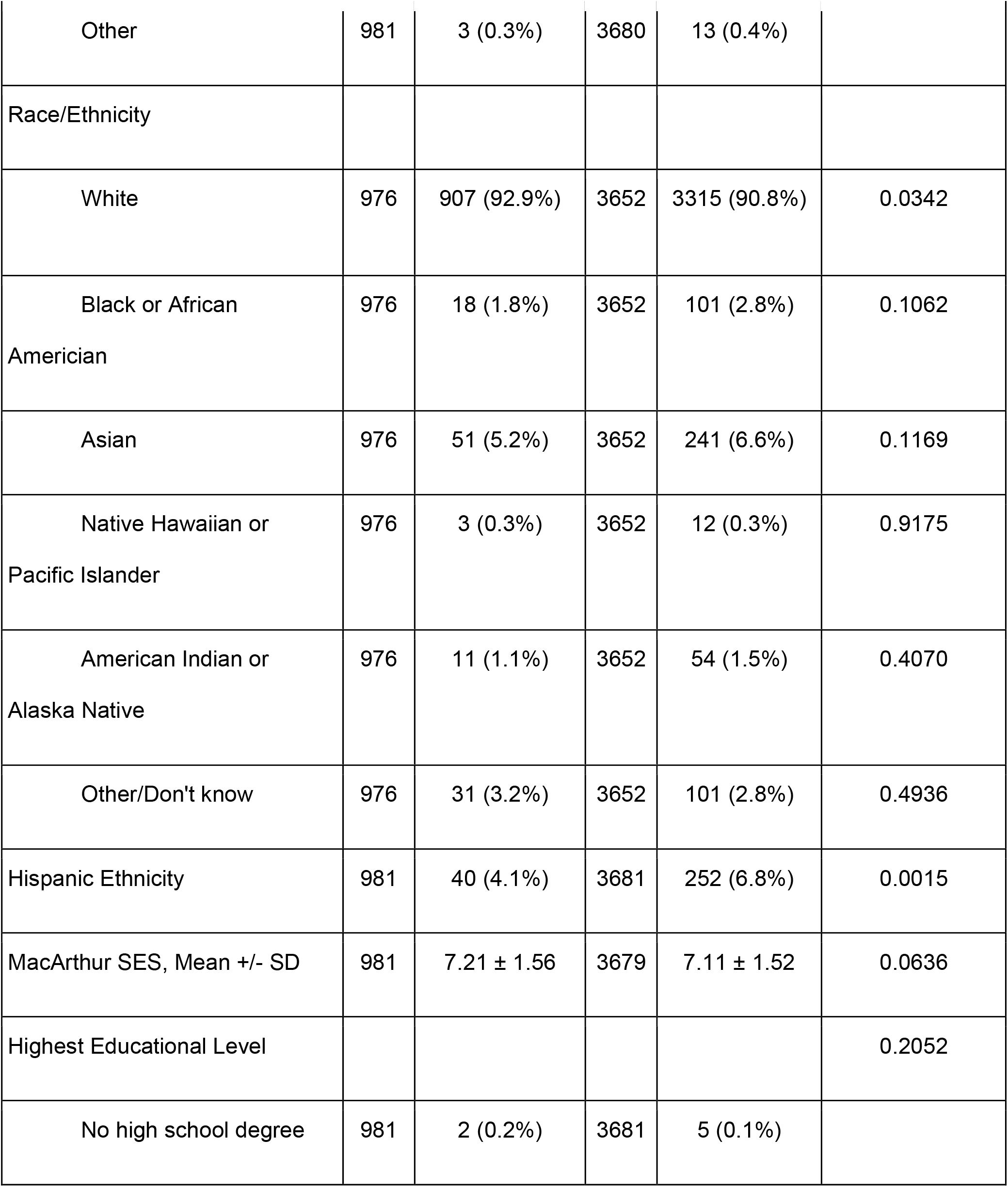
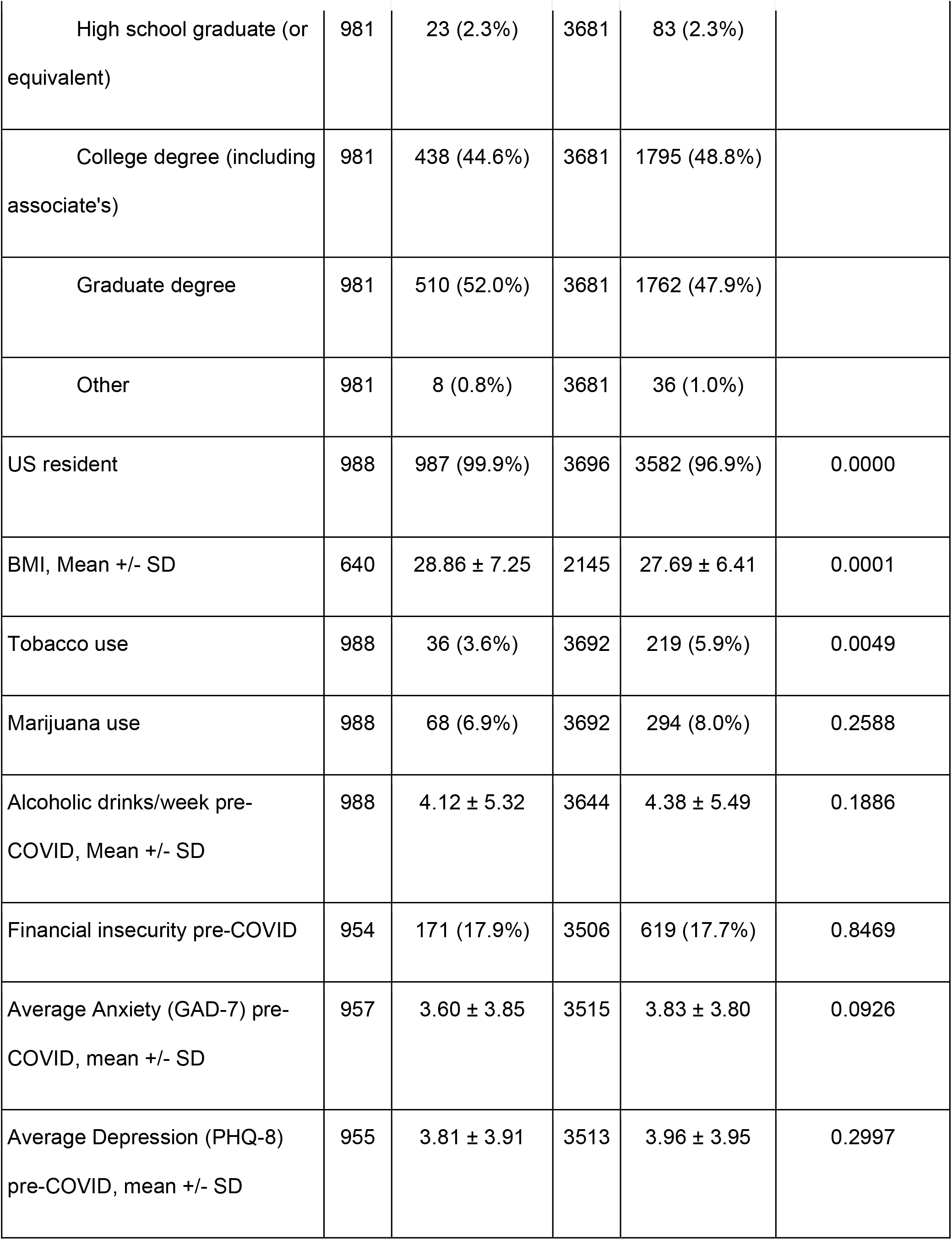
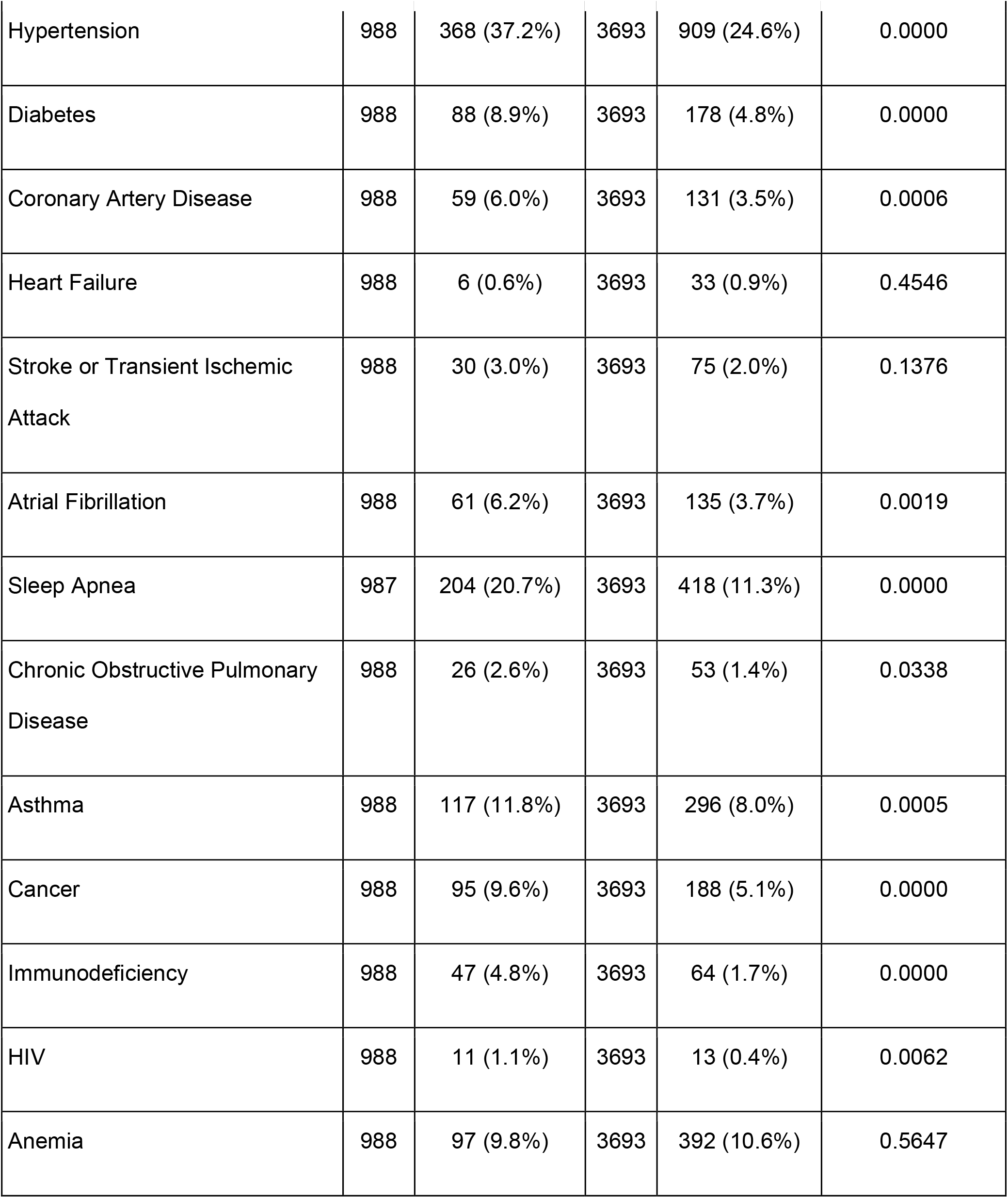
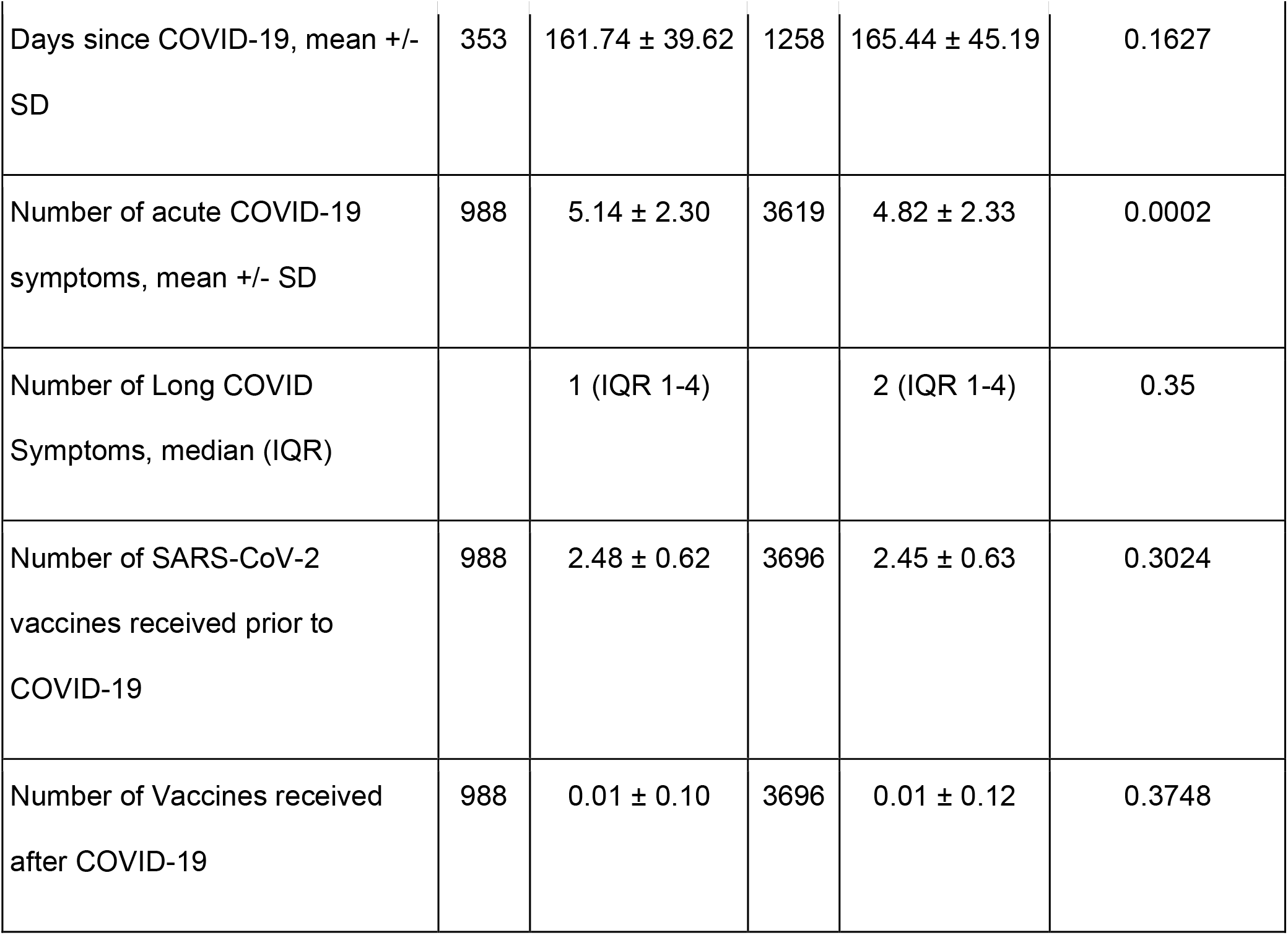
Baseline Demographics and Medical History

|  | **Treated with**<br>**nirmatrelvir/ritonavir,**<br>**N = 988** |  | **SARS-CoV-2**<br>**infected, untreated,**<br>**N = 3696** |  |  |
| --- | --- | --- | --- | --- | --- |
| Characteristic | N | Mean ± std or<br>n(%) | N | Mean ± std or<br>n(%) | P value treated<br>vs not |
| Age, Mean +/- SD | 984 | 59.84 ± 13.30 | 3678 | 52.10 ± 14.02 | 0.0000 |
| Female sex assigned at birth | 981 | 604 (61.6%) | 3681 | 2472 (67.2%) | 0.0010 |
| Gender identity |  |  |  |  | 0.0008 |
| Male | 981 | 376 (38.3%) | 3680 | 1196 (32.5%) |  |
| Female | 981 | 592 (60.3%) | 3680 | 2449 (66.5%) |  |
| Transgender Woman<br>(MTF) |  |  | 3680 | 6 (0.2%) |  |
| Transgender Man (FTM) | 981 | 3 (0.3%) | 3680 | 3 (0.1%) |  |
| Genderqueer | 981 | 10 (1.0%) | 3680 | 16 (0.4%) |  |
| Other | 981 | 3 (0.3%) | 3680 | 13 (0.4%) |  |
| Race/Ethnicity |  |  |  |  |  |
| White | 976 | 907 (92.9%) | 3652 | 3315 (90.8%) | 0.0342 |
| Black or African<br>American | 976 | 18 (1.8%) | 3652 | 101 (2.8%) | 0.1062 |
| Asian | 976 | 51 (5.2%) | 3652 | 241 (6.6%) | 0.1169 |
| Native Hawaiian or<br>Pacific Islander | 976 | 3 (0.3%) | 3652 | 12 (0.3%) | 0.9175 |
| American Indian or<br>Alaska Native | 976 | 11 (1.1%) | 3652 | 54 (1.5%) | 0.4070 |
| Other/Don't know | 976 | 31 (3.2%) | 3652 | 101 (2.8%) | 0.4936 |
| Hispanic Ethnicity | 981 | 40 (4.1%) | 3681 | 252 (6.8%) | 0.0015 |
| MacArthur SES, Mean +/- SD | 981 | 7.21 ± 1.56 | 3679 | 7.11 ± 1.52 | 0.0636 |
| Highest Educational Level |  |  |  |  | 0.2052 |
| No high school degree | 981 | 2 (0.2%) | 3681 | 5 (0.1%) |  |
| High school graduate (or equivalent) | 981 | 23 (2.3%) | 3681 | 83 (2.3%) |  |
| College degree (including associate's) | 981 | 438 (44.6%) | 3681 | 1795 (48.8%) |  |
| Graduate degree | 981 | 510 (52.0%) | 3681 | 1762 (47.9%) |  |
| Other | 981 | 8 (0.8%) | 3681 | 36 (1.0%) |  |
| US resident | 988 | 987 (99.9%) | 3696 | 3582 (96.9%) | 0.0000 |
| BMI, Mean +/- SD | 640 | 28.86 ± 7.25 | 2145 | 27.69 ± 6.41 | 0.0001 |
| Tobacco use | 988 | 36 (3.6%) | 3692 | 219 (5.9%) | 0.0049 |
| Marijuana use | 988 | 68 (6.9%) | 3692 | 294 (8.0%) | 0.2588 |
| Alcoholic drinks/week pre-COVID, Mean +/- SD | 988 | 4.12 ± 5.32 | 3644 | 4.38 ± 5.49 | 0.1886 |
| Financial insecurity pre-COVID | 954 | 171 (17.9%) | 3506 | 619 (17.7%) | 0.8469 |
| Average Anxiety (GAD-7) pre-COVID, mean +/- SD | 957 | 3.60 ± 3.85 | 3515 | 3.83 ± 3.80 | 0.0926 |
| Average Depression (PHQ-8) pre-COVID, mean +/- SD | 955 | 3.81 ± 3.91 | 3513 | 3.96 ± 3.95 | 0.2997 |
| Hypertension | 988 | 368 (37.2%) | 3693 | 909 (24.6%) | 0.0000 |
| Diabetes | 988 | 88 (8.9%) | 3693 | 178 (4.8%) | 0.0000 |
| Coronary Artery Disease | 988 | 59 (6.0%) | 3693 | 131 (3.5%) | 0.0006 |
| Heart Failure | 988 | 6 (0.6%) | 3693 | 33 (0.9%) | 0.4546 |
| Stroke or Transient Ischemic<br>Attack | 988 | 30 (3.0%) | 3693 | 75 (2.0%) | 0.1376 |
| Atrial Fibrillation | 988 | 61 (6.2%) | 3693 | 135 (3.7%) | 0.0019 |
| Sleep Apnea | 987 | 204 (20.7%) | 3693 | 418 (11.3%) | 0.0000 |
| Chronic Obstructive Pulmonary<br>Disease | 988 | 26 (2.6%) | 3693 | 53 (1.4%) | 0.0338 |
| Asthma | 988 | 117 (11.8%) | 3693 | 296 (8.0%) | 0.0005 |
| Cancer | 988 | 95 (9.6%) | 3693 | 188 (5.1%) | 0.0000 |
| Immunodeficiency | 988 | 47 (4.8%) | 3693 | 64 (1.7%) | 0.0000 |
| HIV | 988 | 11 (1.1%) | 3693 | 13 (0.4%) | 0.0062 |
| Anemia | 988 | 97 (9.8%) | 3693 | 392 (10.6%) | 0.5647 |
| Days since COVID-19, mean +/- SD | 353 | 161.74 ± 39.62 | 1258 | 165.44 ± 45.19 | 0.1627 |
| Number of acute COVID-19 symptoms, mean +/- SD | 988 | 5.14 ± 2.30 | 3619 | 4.82 ± 2.33 | 0.0002 |
| Number of Long COVID Symptoms, median (IQR) |  | 1 (IQR 1-4) |  | 2 (IQR 1-4) | 0.35 |
| Number of SARS-CoV-2 vaccines received prior to COVID-19 | 988 | 2.48 ± 0.62 | 3696 | 2.45 ± 0.63 | 0.3024 |
| Number of Vaccines received after COVID-19 | 988 | 0.01 ± 0.10 | 3696 | 0.01 ± 0.12 | 0.3748 |

**Table 2:** Unadjusted Outcomes among Survey Respondents (n=1611)

|  | **Nirmatrelvir/**<br>**ritonavir treated** | **Not treated** | **Total** |
| --- | --- | --- | --- |
| ≥1 Long COVID symptom | 57 (24.5%) | 296 (21.5%) | 353 (21.9%) |
| No Long COVID symptoms | 176 (78.5%) | 1082 (75.5 %) | 1258 (78.1%) |
| Total | 233 | 1378 | 1611 |

### Association of nirmatrelvir/ritonavir treatment with Long COVID symptoms

Among those treated with nirmatrelvir/ritonavir who responded to the Long COVID survey, 57/353 (16.1%) reported Long COVID symptoms compared to 176/1258 (14.0%) who were not treated (OR 1.18; 95%CI 0.84-1.65; p=0.31). As shown in Figure 2, in the propensity-adjusted model, treatment with nirmatrelvir/ritonavir among vaccinated, non-hospitalized individuals was not associated with self-reported Long COVID symptoms (OR 1.15; 95%CI 0.80-1.64; p=0.45). Results were similar when adding race/ethnicity, past medical history, and substance use to the model (OR 1.17; 95%CI 0.81-1.69). Similarly, using inverse probability of treatment weighting as an alternative analytic strategy yielded the same estimate (OR 1.17; 95%CI 0.80-1.71; p=0.43).

**Figure 2.**
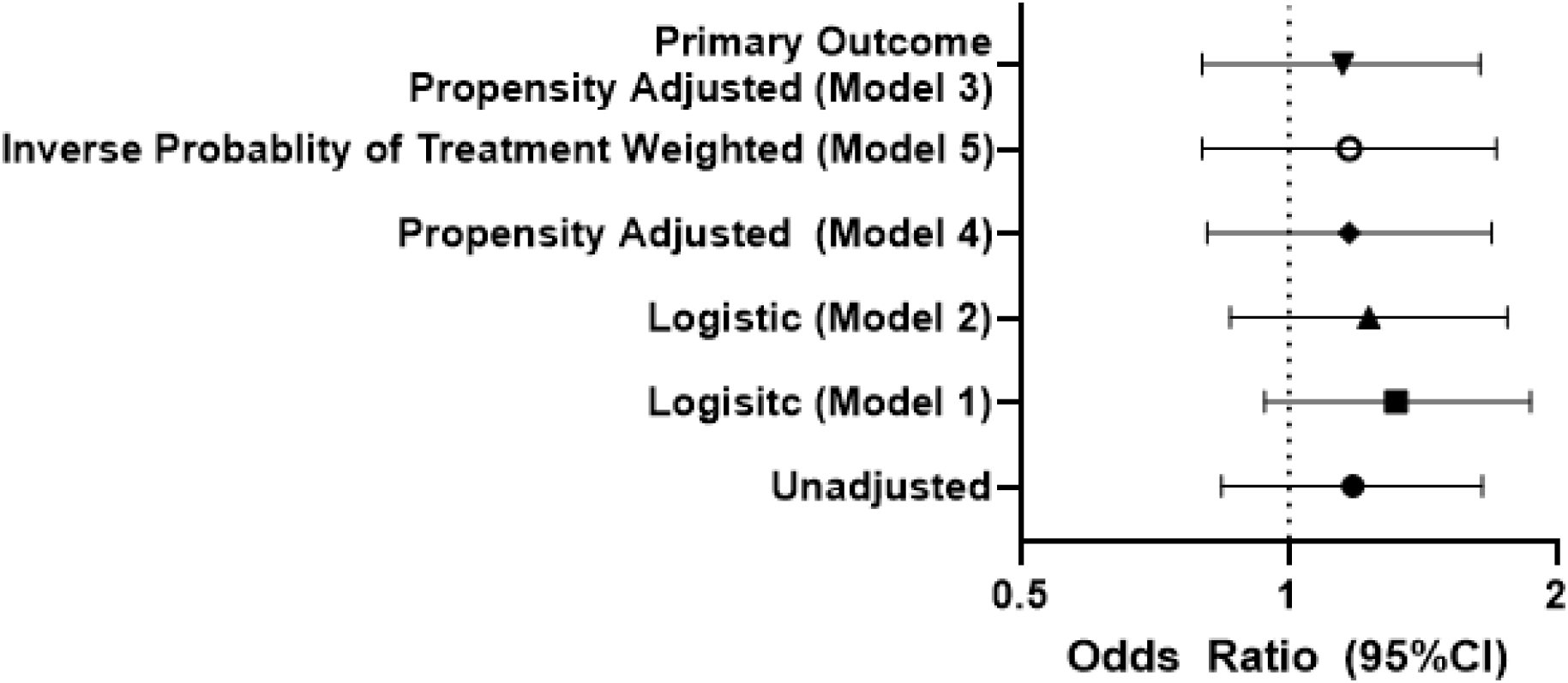
Forest Plot of Treatment Effect of Oral Nirmatrelvir/ritonavir on Long COVID symptoms. In all models, treatment with oral nirmatrelvir/ritonavir was associated with higher odds of patient reported Long COVID symptoms with confidence intervals that cross 1, which suggests that treatment is not beneficial in reducing the risk of Long COVID. Model 3, the pre-specified primary result, includes age, sex, and time since SARS-CoV-2 infection, and the restricted cubic spline of the propensity score. Model 1 includes age, sex, and time since SARS-CoV-2 infection. Model 2 includes age, sex, time since SARS-CoV-2 infection, race/ethnicity, past medical history and substance use. Model 4 is the propensity-adjusted version of Model 2. Finally, Model 5 incorporates inverse probability of treatment weighting and age, sex, and time since SARS-CoV-2 infection.

Among those treated with nirmatrelvir/ritonavir reporting at least one Long COVID symptom, the median number of Long COVID symptoms was 1 (IQR 1-4) compared to 2 (IQR 1-4) among those untreated (p=0.35); the mean number of symptoms was 2.86±2.42 and 3.06±2.54, respectively (p=0.70). Few individuals reported at least one severe or very severe Long COVID symptom in either group: 6 (1.7% overall, 10.5% with Long COVID) among the treated and 18 (1.4% overall, 10.2% with Long COVID) among the untreated (p=0.18); because few participants reported many or severe symptoms, we could not exclude an effect on symptom severity.

### Rebound

Among 666 individuals who responded to the rebound survey at least 30 days after taking nirmatrelvir/ritonavir for acute SARS-CoV-2 infection, 139/650 (21%) reported rebound symptoms. Among those who repeated antigen testing after completing treatment, 97/377 (25.7%) reported rebound test positivity. In total, 174/666 (26.1%) experienced rebound symptoms or test positivity. Among those with rebound, 18/166 (10.8%) reported 1 or more Long COVID symptom compared to 39/468 (8.3%) who did not experience rebound (OR 1.34; 95%CI 0.74-2.41; p=0.33). Results were similar when considering rebound symptoms (9.9% vs 8.4%; OR 1.20; 95%CI 0.62-2.31; p=0.59) or rebound test positivity (10.8% vs 7.7%; OR 1.45; 95%CI 0.66-3.21; p=0.36). Results were similar using a more sensitive outcome of those who had not experienced full recovery (14.9% with rebound vs 12.8% without rebound; OR 1.20; 95%CI 0.73-1.96; p=0.48).

Results were similar when limiting analysis to respondents who completed treatment more than 90 days prior (n=157). Of those, 31/157 (20.2%) experienced rebound symptoms and 17/84 (19.7%) had a rebound test positivity with 38/157 (24.2%) classified as having any rebound. Only 2/38 (5.2%) individuals who experienced rebound symptoms reported Long COVID symptoms compared to 19/119 (16.0%) who did not experience rebound (p=0.11). No individuals who experienced rebound test positivity reported Long COVID symptoms (0/17) compared to 10/67 (14.9%) among those with repeated testing without rebound test positivity (p=0.20).

## Discussion

Within an online observational cohort, we did not find evidence that treatment with oral nirmatrelvir/ritonavir during the acute phase of COVID-19 was associated with a lower prevalence of patient-reported Long COVID symptoms at least 90 days after infection among vaccinated, non-hospitalized individuals experiencing their first known SARS-CoV-2 infection. We did not find a significant effect of treatment on the number of Long COVID symptoms or severe symptoms, although these endpoints were limited by a small number of people experiencing more than 1 symptom or severe symptoms. Similarly, we did not find that rebound symptoms or test positivity after treatment with nirmatrelvir/ritonavir were associated with a statistically significant effect on Long COVID symptoms.

### Prior studies of nimatrelvir/ritonavir focus on acute symptoms

Prior studies of oral nimatrelvir/ritonavir, including two randomized clinical trials, EPIC-HR and EPIC-SR, have focused on acute outcomes of SARS-CoV-2 infection. EPIC-HR demonstrated a reduction in hospitalization and mortality by day 28 among those at high risk of disease progression treated with nirmatrelvir/ritonavir compared to placebo [9], but the EPIC-SR study of standard risk individuals was stopped early after there was no benefit in improvement of symptoms or progression to severe disease [18]. A number of “real-world” observational studies have mostly found similar results, with reductions in hospitalization and mortality among higher risk individuals as well as vaccinated individuals [19-23]. Others have demonstrated faster viral clearance with nirmatrelvir/ritonavir [24, 25]; as higher viral loads and prolonged viral shedding have been previously associated with risk of Long COVID [3, 4], there is a strong rationale for the hypothesis that treatment with nirmatrelvir/ritonavir may lower the risk of Long COVID. Secondly, in this cohort, we have previously shown that the number of symptoms during acute infection is associated with Long COVID symptoms independent of vaccination and variant wave [15], but whether reducing acute symptoms with antiviral therapy reduces risk of Long COVID has not been demonstrated.

### Our study in context of studies of nirmatrelvir/ritonavir on post-COVID outcomes

Only two [yet unpublished] studies have considered the potential impact of oral nirmatrelvir/ritonavir at the time of acute infection on longer term post-COVID outcomes, with contradictory results despite both including Veterans and examining EHR-diagnosed post-COVID conditions by ICD-10 diagnostic codes as the outcome of interest [13, 26]. Bajema et al found a benefit on acute outcomes within 30 days but did not find a reduction in EHR-diagnosed post-COVID conditions among the treated at 6 months, whereas Xie et al found a decrease in post-COVID conditions, hospitalizations, and mortality among treated individuals from 30-90 days. One possible explanation for the difference is that Bajema et al used a target-trial emulation study design, which is more robust to common pitfalls of observational studies of treatment effects. Both studies focused on an older, sicker patient population and are limited by differential outcome ascertainment, differential misclassification, and lack of ascertainment that these ICD-codes correspond to patient symptoms.

In contrast to EHR-based studies focused on ascertainment of post-COVID-conditions using ICD-10 codes, our study evaluated whether treatment with oral nimatrelvir/ritonavir has an impact on patient-reported Long COVID symptoms, as well as the association between patient-reported rebound and subsequent Long COVID symptoms. Our outcome of patient-reported symptoms is a closer approximation of the condition of interest (Long COVID, defined as unexplained symptoms persisting for >90 days following initial infection in accordance with the WHO definition) [1]. Our finding that nimatrelvir/ritonavir treatment during acute infection is not associated with lower odds of Long COVID is consistent with the Bajema et al report from the VA that there was no difference in post-COVID conditions at 6 months [13]. Data on the effect of other acute therapies including remdesivir on Long COVID symptoms have been mixed, with Italian observational data suggesting a benefit with remdesivir [27], but the Finish randomized SOLIDARITY trial did not demonstrate a difference in Long COVID with remdesivir compared to placebo [28]. One study suggested a benefit from metformin but not ivermectin or fluvoxamine [29], but a randomized platform study did not find that doxazosin, fluvoxamine, fluvoxamine in combination with inhaled budesonide, interferon-lambda, ivermectin, or metformin had any impact on Long COVID symptoms or quality of life [30].

### Rebound and Long COVID

We found a higher proportion with clinical rebound than reported in the literature [31], but did not identify an effect of post-treatment rebound on the later development of Long COVID symptoms. Since antivirals entered widespread use, there has been intense interest in this phenomenon, and some case reports have clearly demonstrated the development of Long COVID among those with post-treatment rebound [10]. The observation that anti-viral associated rebound is not associated with Long COVID in this study suggests that the potential for treatment-related rebound symptoms should not discourage antiviral use in the first place, and that individuals experiencing this phenomenon are not uniquely at risk for Long COVID. However, rebound test positivity was common in the early days of the pandemic [32]; because of the way our data were collected, we were not able to determine whether the presence of rebound unrelated to treatment increases the likelihood of an individual experiencing Long COVID. If this were the case, it is possible that rebound symptoms could be used as a prognostic marker of later post-acute symptoms.

### Role of treatment for Long COVID

Although a 5 day course of oral nimatrelvir/ritonavir at the time of acute test positivity in this low-risk, vaccinated population was not associated with a reduction in Long COVID, our study does not consider whether nimatrelvir/ritonavir may be effective at treating Long COVID, which is currently under investigation in one ongoing and two planned clinical trials (NCT05576662, NCT05595369, NCT05668091).

### Limitations

The primary limitations of our study arise from its observational nature. The cohort was relatively homogeneous; most individuals identified as white with advanced education, limiting the generalizability of the findings, in large part because we limited this study to first infections and thus only those privileged enough to avoid infection until March 2022 were included. We relied on participant self-report of nirmatrelvir/ritonavir treatment and Long COVID symptoms. SARS-CoV-2 testing and nirmatrelvir/ritonavir use were collected longitudinally at the time of positive test and treatment, respectively, limiting the role of recall bias in ascertaining exposure. We did not include the number of symptoms during acute infection in our models as we could not determine their temporal relation to treatment. Participant self-report of Long COVID symptoms is currently the gold-standard for ascertainment of the outcome. The survey response rate was non-differential between treated and untreated individuals. We note that our study differs from EHR-based reports in that we assessed Long COVID symptoms rather than the presence of PASC as measured through clinician-coded ICD-10 codes. We therefore could not determine whether treatment during the acute phase had an impact on other important post-acute outcomes like cardiovascular sequelae, diabetes, or other medical diagnoses. We also could not ascertain whether treatment had an effect on objectively measured post-COVID outcomes (e.g., exercise capacity [33], neurocognitive performance [34], or other objectively measurable physiologic perturbations) potentially associated with Long COVID which may lend additional specificity to the outcome. We used propensity scores and inverse probability of treatment weighting to adjust for baseline differences between propensity of treatment between the treated and untreated, but residual confounding may still bias the results. An additional limitation regarding the rebound test positivity analysis is that tests were not performed systematically, and results were based on participant test self-report. We did not assess for rebound symptoms among those not treated.

## Conclusions

Among vaccinated, non-hospitalized adult participants in the COVID Citizen Science online cohort, we did not find that treatment with oral nirmatrelvir/ritonavir during the acute phase of SARS-CoV-2 infection was associated with lower odds of Long COVID symptoms more than 90 days after index infection. Among those treated, we did not find that rebound was associated with a large increase in Long COVID symptoms.

## Data Availability

All data produced in the present study are available upon reasonable request to the COVID Citizen Science Steering Committee.

## Acknowledgements

We would like to thank all of the research participants and all of the Patient-Centered Outcomes Research Institute sites and investigators who recruited participants.

